# Low-Dose Whole-Lung Radiation for COVID-19 Pneumonia: Planned Day-7 Interim Analysis of an Ongoing Clinical Trial

**DOI:** 10.1101/2020.06.03.20116988

**Authors:** Clayton B. Hess, Zachary S. Buchwald, William Stokes, Jeffrey M. Switchenko, Tahseen H. Nasti, Brent D. Weinberg, James P. Steinberg, Karen D. Godette, David Murphy, Rafi Ahmed, Walter J. Curran, Mohammad K. Khan

## Abstract

**Background:** Individuals with advanced age and comorbidities face higher risk of death from COVID-19, especially once ventilator-dependent. Respiratory decline in COVID-19 is mediated by a pneumonic aberrant immune cytokine storm. Low-dose radiation was used to treat pneumonia in the pre-antibiotic era. Radiation immunomodulatory effects may improve outcomes in COVID-19.

**Methods:** We performed a single-institution phase I/II trial evaluating the safety and efficacy of single-fraction, low-dose, whole-lung radiation for COVID-19 pneumonia. Eligible patients were hospitalized, had radiographic pneumonic infiltrates, required supplemental oxygen, and were clinically deteriorating.

**Results:** Of nine patients screened, five were treated with whole-lung radiation from April 23-28, 2020 and followed for 7 days. Median age was 90 (range 64-94); four were nursing home residents with multiple comorbidities. Within 24 hours of radiation, three patients (60%) weaned from supplemental oxygen to ambient air, four (80%) exhibited radiographic improvement, and median Glasgow coma score improved from 10 to 14. A fourth patient (80% overall recovery) weaned from oxygen at hour 96. Mean time to clinical recovery was 35 hours. There were no acute skin, pulmonary, GI, GU toxicities.

**Conclusions:** In a small pilot trial of five oxygen-dependent patients with COVID-19 pneumonia, low-dose whole-lung radiation led to rapid improvement in clinical status, encephalopathy, and radiographic infiltrates without acute toxicity or worsening the cytokine storm. Low-dose whole-lung radiation appears to be safe, shows early promise of efficacy, and warrants larger prospective trials.

**Lay Summary:** Researchers at Emory University have completed treatment of cohort 1 of a pilot trial of low-dose lung irradiation for COVID-19 pneumonia. Five residents of nursing or group homes with COVID-19 outbreaks were hospitalized after testing positive for COVID-19. Each had pneumonia visible on chest x-ray, required supplemental oxygen, and had clinically declined. A single treatment of low-dose (1.5 Gy) radiation to both lungs was delivered over 10-15 minutes. Within 24 hours, four patients rapidly improved their breathing, recovering at an average of 1.5 days (range 3 to 96 hours), and had either been discharged (three) or were preparing for discharge (one) by day 14. Blood tests and repeat imaging confirmed that low-dose radiation appeared safe and effective in reducing their COVID-19 symptoms. Further trials are warranted.

## Introduction

Individuals with advanced age and comorbidities face a higher risk of death from COVID-19, especially once ventilator-dependent, precipitated by an immune cytokine storm in the lungs.^1^’^3^ Lymphocytes, a mediator of cytokine storms, are exquisitely sensitive to ionizing radiation.^4^ Low doses of radiation therapy (LD-RT) were successfully used to treat patients with infectious processes during the first half of the 20th century, including pneumonia.^5^ It is conceivable that focal immunosuppression with LD-RT may reduce pulmonary inflammation associated with COVID-19 pneumonia and thereby alter the clinical course of infected patients.

## Methods

### Trial Design

The Radiation Eliminates Storming Cytokines and Unchecked Edema as a 1-day Treatment for COVID-19 (RESCUE 1-19) trial is an ongoing investigator-initiated trial exploring safety and efficacy of single-fraction, low-dose, whole-lung radiation for hospitalized, oxygen-dependent patients with COVID-19 pneumonia (Clinical Trial Registration Number NCT04366791). The protocol was approved by the Emory University Institutional Review Board, and all human participants or their surrogate representatives gave written informed consent. Eligibility required patients to be hospitalized with a positive nasopharyngeal swab for COVID-19, have radiographic pneumonic consolidations, require oxygen supplementation, and be clinically deteriorating (i.e. mentation, oxygenation, dyspnea).

Exclusion criteria included receipt of any COVID-directed drug therapy within one day before or three days following LD-RT. Intubated patients were deemed unstable for transport and not treated. Patients received a single-fraction radiation dose of 1.5 Gy to the bilateral lungs, delivered via an anteriorposterior beam configuration. The primary endpoint was safety as assessed by clinical, radiographic, and inflammatory marker response. Clinical recovery was defined as the first day a subject was discharged or weaned from supplemental oxygen.

### Results

Between April 23-28, 2020, nine study candidates were evaluated. Of these, one died before enrollment, one did not meet severity criteria, and seven enrolled. Of these, two were intubated before LD-RT (one died), and five were treated with LD-RT. Table 1 outlines demographic and clinical characteristics of treated patients at admission. Median age was 90 (range 64-94). Four had been admitted from nursing homes with COVID-19 outbreaks. Comorbidity burden was high. Four were African American, and one was Caucasian. Four were female. Median Glasgow Coma Scale (GCS) score was 10 (range 9-15) at time of consent. Median Karnofsky Performance Status was 40% (range 40-80%). Median oxygen supplementation requirement at the time of LD-RT was 3 L/min (range 1.5-6). Median duration of pre-hospitalization symptoms was 4 days (range 1-7). LD-RT was delivered on median hospital day 5 (range 2-8).

**Table 1.**
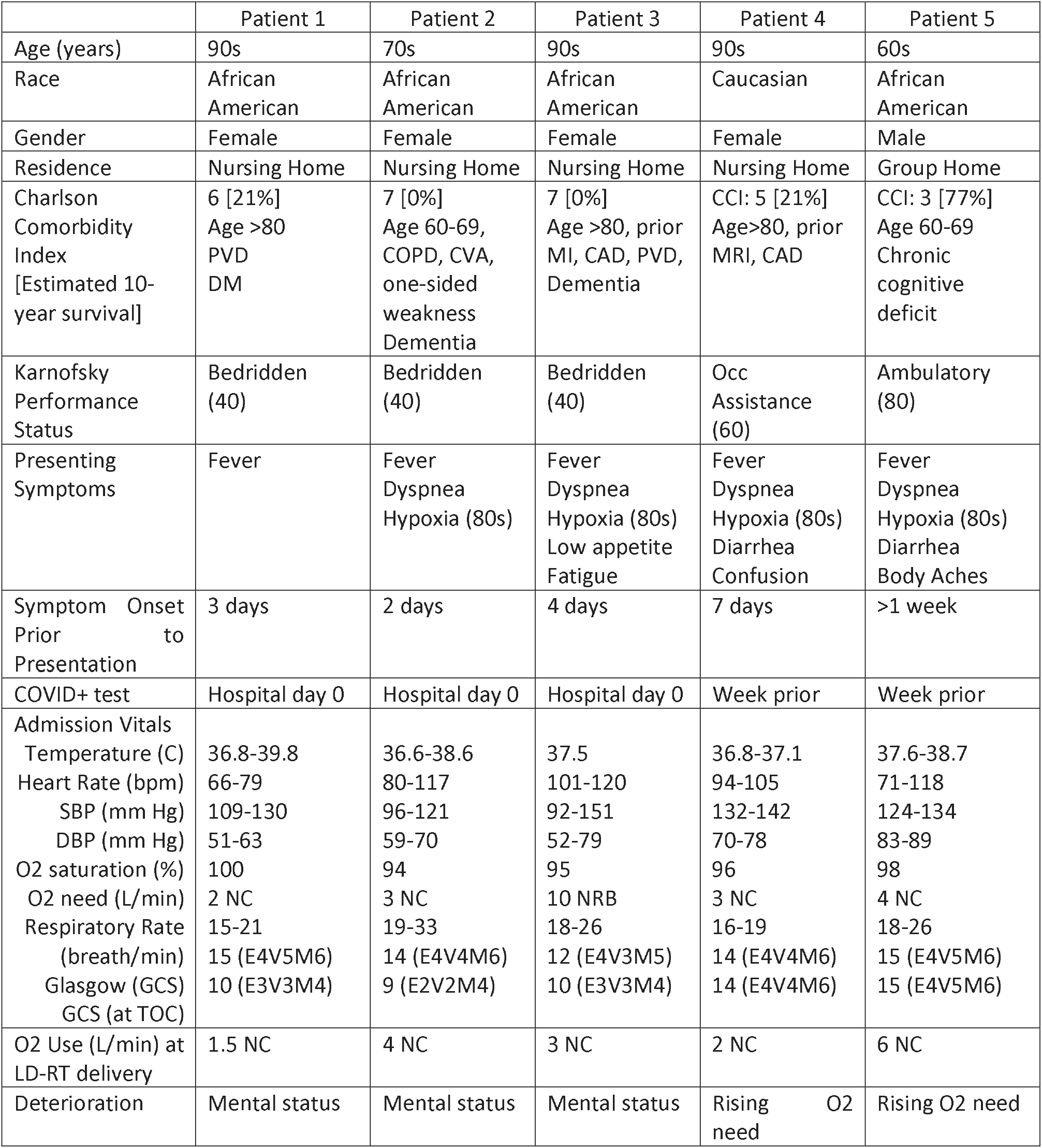
Patient Demographic and Clinical Characteristics at Time of Admission

During a 14-day observation period, 4 patients (80%) clinically recovered, 3 within 24 hours (Figure 1). Mean time to recovery was 1.5 days (35 hours). Mean time to discharge was 12 days for the 4 patients. Median GCS rose from 10 (range 9-15) to 14 (range 13-15) at hour 24. No acute skin, GI, cardiac, or pulmonary toxicities were noted. At day 3, 80% of assessed inflammatory biomarkers either remained stable or improved. At day 7, all but one patient with abnormal inflammatory (4/5), renal (2/3), cardiac (4/5), clotting (3/4), and hepatic (3/4) blood markers had rapidly returned to or were approaching normal levels. At day 14, all patients were alive, 3 had returned to their nursing homes, and a 4th was pending hospital discharge.

**Figure 1.**
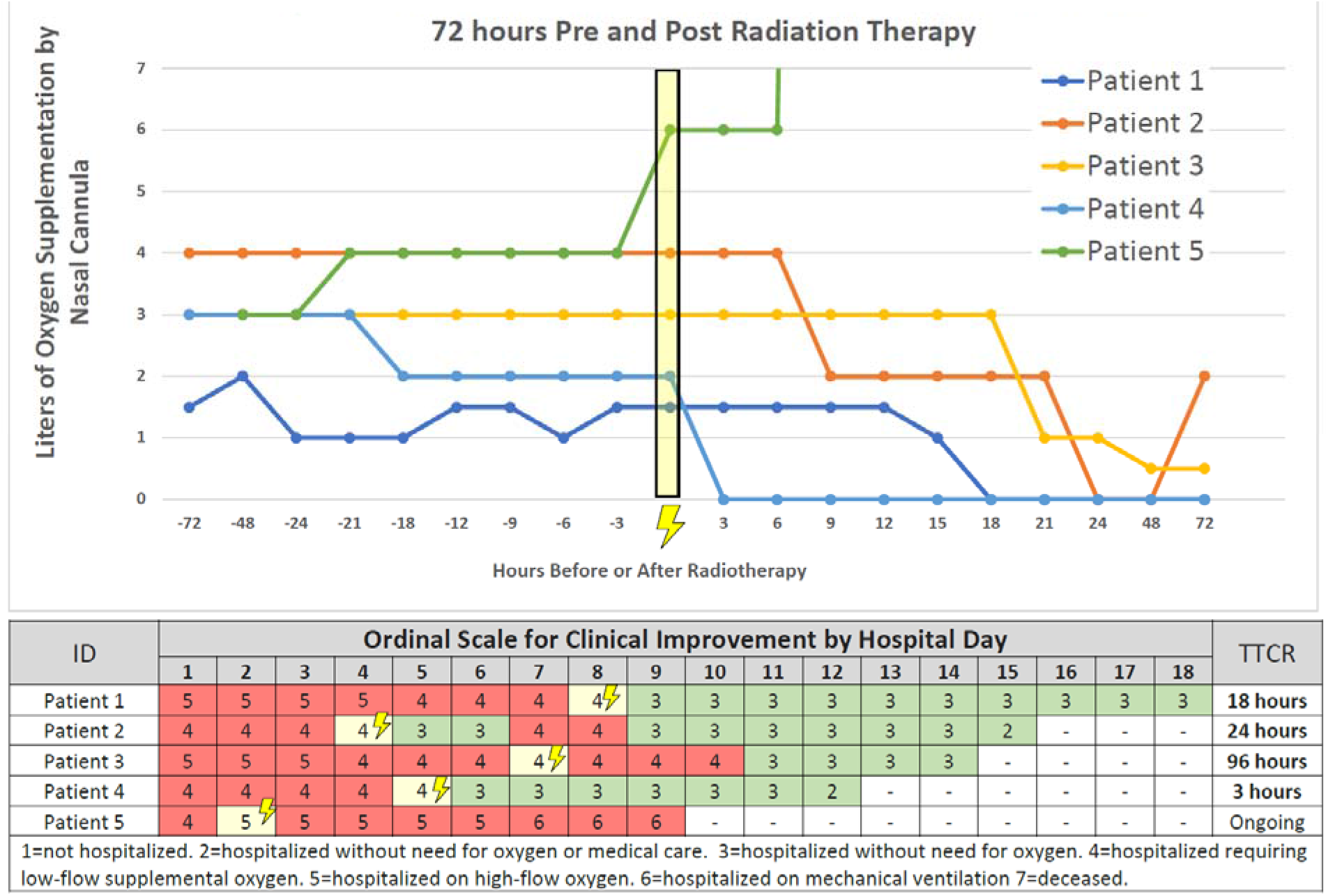
Time to Clinical Recovery (TTCR) and Oxygen Saturations 72 hours Pre and Post Radiation Therapy

## Discussion

This letter summarizes the first known use of ionizing radiation for infectious disease in the modern era and the first use in world, to author’s knowledge, for patients with COVID-19. Five hospitalized, oxygen-dependent, and clinically deteriorating patients received low-dose, whole-lung radiation and experienced no acute toxicity. 80% of patients returned to breathing room air at a median time of 1.5 days. Two patients who consented to the trial rapidly deteriorated and could not receive LD-RT, yielding a cumulative response rate for the entire cohort of 4/7 (57%). No toxicities, including dermatitis, cytopenia, or worsening cytokine storm, were observed. Low-dose lung radiation appears safe and merits further evaluation to determine efficacy among patients with COVID-19 pneumonia.

## Data Availability

All data is included in the manuscript that has been uploaded to the server.

## Acknowledgements

We would like to acknowledge our collaborating physicians Dr. Jesse James MD, Dr. Michael Sterling MD, Dr. Charles Grodzin MD, Dr. Craig Coopersmith MD, Dr. Greg Martin MD, Dr. Marybeth Sexton MD MSC, Dr. Ramzy Rimawi MD, and Dr. Samer Melhem MD PhD for contributions and collaboration for trial accrual.

